# A Novel FiO2 Titration Protocol for Quantifying Pulmonary Oxygen Reserve Capacity : Dynamic Assessment Framework for Infection-Associated Respiratory Dysfunction

**DOI:** 10.1101/2025.04.21.25326055

**Authors:** Shi Qiru

## Abstract

2

**Background:** Progressive decline in pulmonary oxygen reserve capacity (ORC) is a hallmark of infection-associated respiratory dysfunction. Current tools (PaO2/FiO2 ratio, cardiopulmonary exercise testing [CPET], computed tomography [CT]) are limited in dynamic monitoring due to delayed responsiveness, operational complexity, or radiation risks, and other constraints.

**Methods:** The ORC testing methodology integrates the dynamic load-incrementation logic of cardiopulmonary exercise testing (CPET) with the oxygenation quantification framework of PaO2/FiO2. Its operational paradigm comprises three phases:

➀Testing Protocol

Conducted under ventilation-locked conditions, a stepwise FiO2 titration protocol is applied, with termination triggered when SpO2

➁Parameter Definition

The minimum FiO2 required to maintain SpO2 ≥90% (FiO2-MIN) is recorded, and the oxygen reserve capacity is calculated as ORC = 0.21 - FiO2-MIN.

➂Dynamic Modeling

Through continuous monitoring throughout the entire disease course, ORC time-series data are acquired. A time-ORC curve is then fitted, and based on differential calculus (β = ΔORC/Δt, γ = Δβ/Δt), they collectively establish a quantitative respiratory compensation dynamics model in conjunction with the time-ORC curve.

**Results:** The ORC test provides a novel non-invasive tool for dynamic quantification of respiratory reserve. Early warning of ARDS transformation during acute infection and quantitative dynamic tracking of lung dysfunction of long-COVID syndrome are its potential application scenarios. Its clinical utility requires prospective validation through multicenter trials integrated with CPET and CT quantitative analysis.

## 3. Introduction

### 3.1 Limitations of Current Theories

Infection-related respiratory dysfunction (e.g., COVID-19, influenza pneumonia) often involves progressive decline in pulmonary oxygen reserve capacity (ORC). However, existing assessment tools face significant limitations in dynamic monitoring and early warning:

### 3.1.1 Functional Compensatory Blind Zone of the PaO2/FiO2 Ratio

The PaO2/FiO2 ratio, a widely utilized clinical marker of oxygenation, exhibits functional compensatory blind zones during disease progression. Specifically, in the compensatory phase of respiratory dysfunction, patients may maintain a relatively stable PaO2/FiO2 ratio despite significant reductions in pulmonary oxygen reserve capacity (ORC). This discrepancy arises because the ratio fails to reflect dynamic decay in lung reserve, leading to delayed clinical intervention.

### 3.1.2 Practical Constraints of Cardiopulmonary Exercise Testing (CPET)

For chronic pulmonary dysfunction (e.g., long-COVID syndrome), CPET can identify reserve abnormalities such as reduced oxygen uptake (VO2 peak)[1]. However, its accuracy and utility for longitudinal tracking are limited by patient compliance, operational standardization challenges, and equipment dependency.

### 3.1.3 Limitations of computed tomography [CT]

Three-Dimensional Computed Tomography (3D-CT) provides critical quantitative analysis for pulmonary diseases such as COVID-19, enabling clinicians to assess disease severity, monitor progression, and evaluate therapeutic efficacy with enhanced precision. However, this approach faces technical and methodological limitations, including challenges in lung segmentation algorithms, as well as clinical applicability constraints such as observer variability in interpreting heterogeneous lesion patterns.[2]

### 3.2 Theoretical Innovations

The ORC test integrates principles from CPET (dynamic load escalation) and the PaO2/FiO2 framework (oxygenation quantification) while achieving paradigm shifts:

#### 3.2.1 Inheritance and Transformation of CPET Principles

Cardiopulmonary Exercise Testing (CPET) directly quantifies systemic oxygen metabolism reserve capacity to a certain extent by measuring VO2 peak under incremental exercise workloads. However, its results are confounded by exercise-induced ventilatory compensation (e.g., surges in respiratory rate and tidal volume), which obscures the distinction between pulmonary oxygen metabolic reserve and ventilatory compensatory contributions. Moreover, it is influenced by multiple factors such as subject tolerance, leading to difficulties in accurately assessing oxygenation reserve capacity. [3]The Oxygen Reserve Capacity (ORC) test proposed in this study inherits the core principle of “metabolic load stimulation” while achieving dual transformative innovations:

##### ➀ Transformation of Load Modality

Replaces exercise-based workloads with stepwise FiO2 reduction (progressively decreasing inhaled oxygen concentration) to simulate escalating metabolic demand. Measures the pulmonary oxygen compensatory limit via ORC = 0.21 − FiO2-MIN, indirectly quantifying oxygen reserve capacity while circumventing CPET’s exercise dependency and cardiopulmonary risks.

##### ➁Transformation of Functional Analysis

Locks ventilatory parameters (standardized tidal volume and respiratory rate) to eliminate interference from ventilatory compensation on oxygenation signals.

Focuses solely on parenchymal oxygen exchange and microcirculatory regulation, overcoming the mechanistic ambiguity of “ventilation-metabolism coupling” in CPET. Enables precise decoupled evaluation of pulmonary oxygen metabolic reserve. 3.2.2Expansion of the PaO2/FiO2 Framework

##### ➀Assessment Logic Shift

Moves from passive compensation (raising FiO2 to maintain PaO2) to active reserve probing (lowering FiO2 to unmask thresholds). Locked ventilation parameters ensure thresholds reflect intrinsic pulmonary function, not ventilatory variability.

##### ➁Dynamic Quantification

Transforms PaO2/FiO2 from a static snapshot into a longitudinal reserve parameter (ORC) with kinetic derivatives (β = ΔORC/Δt, γ = Δβ/Δt). These metrics reveal reserve decay rates and critical inflection points.

### 3.3 Research Objectives

This study aims to establish a dynamic ORC assessment framework using stepwise FiO2 titration. Specific goals include:

#### ➀Establish a theoretical framework

Propose a stepwise FiO2 reduction-driven dynamic variation model of pulmonary oxygen reserve capacity based on the principle of metabolic load simulation.

#### ➁Standardized ORC Testing Protocol

Operational guidelines: FiO2 titration rate, safety thresholds (e.g., SpO2

#### ➂ORC-β/γ Kinetic Model

Construct a dynamic assessment framework integrating ORC curves, decay rate (β), and acceleration of decay (γ) to quantify pulmonary reserve deterioration.

## 4. Method Design

### 4.1 Testing Principles

The theoretical core of the stepwise FiO2titration method for quantifying pulmonary oxygen reserve capacity (ORC) liesin resolving the oxygen metabolic compensation limits under standardizedpulmonary ventilation. Its design logic inherits and adapts the “metabolicload challenge” principle from cardiopulmonary exercise testing (CPET),while expanding the static assessment paradigm of the PaO2/FiO2 framework. Thisapproach enables dynamic quantification of oxygenation reserve and mechanisticdecoupling of underlying physiological processes.

#### 4.1.1 Origin of ORC: Clinical PracticalAnalysis of PaO2/FiO2

Critically ill patients require incrementalFiO2 adjustments to maintain SaO2 ≥90%. When the FiO2requirement increases accompanied by a concurrent decline in the oxygenationindex (PaO2/FiO2), it suggests deterioration of pulmonary oxygenation function;conversely, a reduction in FiO2 requirement paired with recovery of theoxygenation index indicates improvement in pulmonary function. [4]This negativecorrelation between FiO2 demand and reserve capacity forms the clinicalrationale for ORC.

#### 4.1.2 Inheritance and Transformation ofCPET Principles

Cardiopulmonary Exercise Testing (CPET)evaluates systemic oxygen metabolism limits by incrementally increasing exerciseloads, following the principle of “metabolic demand driving oxygenationdemand.” Similarly, ORC testing reduces FiO2 (thereby limiting oxygensupply) to force the lung-microcirculation unit to maximize oxygen extractionefficiency under constrained oxygen availability. This process mimics thecompensatory limits of oxygen utilization during increased metabolic demand,analogous to CPET’s ability to elicit VO2 peak under incremental exerciseloads. By standardizing ventilation parameters, ORC testing isolates pulmonaryoxygen metabolism reserve capacity while eliminating confounding factorsassociated with physical activity.

#### 4.1.3 Definition of ORC and TheoreticalValidation

##### ➀ ORC Test Protocol

Under standardized ventilation (fixed VTand RR, sitting position), FiO2 is stepwise reduced (0.01 decrements) whilemonitoring SpO2. The minimum FiO2 required to sustain SpO2 ≥90%(FiO2-MIN) is recorded.

SpO2 vs. SaO2: SpO2 provides noninvasive,real-time monitoring, avoiding arterial blood sampling while enabling dynamicFiO2 adjustments.[5]

Formula:

ORC=0.21−FiO2-MIN

Theoretical Basis: 0.21 represents ambientair oxygen concentration (21%). FiO2-MIN reflects the minimum oxygen demand tosustain baseline oxygenation. ORC quantifies the “safety margin” ofpulmonary oxygen reserve.

Negative ORC Values: When FiO2-MIN >21%, ORC becomes negative, indicating complete depletion of pulmonary oxygenreserve (i.e., supplemental oxygen is required to maintain SpO2 ≥ 90%).

##### ➁ Consistency with Critical Care Practice

In critically ill patients, rising FiO2-MIN(ORC↓) correlates with worsening oxygen reserve (e.g., decliningPaO2/FiO2), while falling FiO2-MIN (ORC↑) indicatesrecovery. Changes in ORC during the course of critical illness are fullyconsistent with clinical observations of pulmonary oxygenation function,validating its reliability as a quantitative indicator of oxygen reserve.

##### ➂ Applicability to Non-Critical/Early-Stage Patients

Even in non-severe cases, reduced ORC(FiO2-MIN↑) reflects early impairment in oxygen exchange efficiency, sharingpathophysiological mechanisms with critical illness. ORC provides a unifiedmetric for longitudinal monitoring across disease stages.

#### 4.4.4 Physiological Basis for VentilationStandardization

##### ➀ Determinant Role of Alveolar Ventilation

Alveolar ventilation (VA=(VT−VD)×RR, whereVD = dead space volume) must remain stable during ORC testing. Fixing VT and RReliminates ventilation-induced variability in oxygenation, ensuring ORCreflects only pulmonary parenchymal and microcirculatory function [6].

##### ➁ Necessity of Stable Ventilation/Perfusion (V/Q) Ratios

Spontaneous breathing causes dynamicfluctuations in VT/RR (e.g., sighing breaths), leading to V/Q mismatch andconfounding FiO2-MIN measurements. Standardization strategies include:

Spontaneously Breathing Patients: Real-timebiofeedback (e.g., auditory cues and respiratory flow meters) to maintain VTvariability ≤±15%; testing pauses if thresholds are exceeded.

Mechanically Ventilated Patients: PredefinedVT=6 – 8 mL/kg and RR=12 – 20 breaths/min.

##### ➂ Standardization of Testing Posture

Posture impacts respiratory mechanics. Forexample, supine positioning increases abdominal pressure on the diaphragm,reducing functional residual capacity (FRC) and vital capacity (VC) compared toupright positions [7].

Non-Critical/Early-Stage Patients: Seatedposition with trunk upright (90°). Critically ill patients: Semi-recumbentposition (30–45° trunk elevation) to minimize posture-related variability.

##### ➀ Definitions and Calculations

#### 4.1.5 β-γ Kinetic Model

β=ΔORC/Δt(unit: day^-1^)

γ=Δβ/Δt (unit: day^-2^)

##### ➁ Clinical Significance

β < 0: ORC decreases over time, indicating deterioration ofoxygenation reserve.

γ > 0: Acceleration of β (improving recovery rate).

#### 4.1.6 Theoretical Model Validation

##### ➀ Repeatability Verification

Three measurements within 4 hours(excluding acute deterioration phases) must show ORC variability < 5%.

##### ➁ Pathological Correlation

ORC and β-γ values must correlatesignificantly with gold-standard parameters (e.g., diffusing capacity foroxygen, DLO2) and clinical outcomes (e.g., incidence of respiratory failure).

##### ➂ Ethics and Safety Protocol

High-risk patients (e.g., coronary arterydisease, pulmonary hypertension) are excluded. Emergency oxygen rescue protocols aremandated (e.g., instantaneously restoring FiO2 to 0.5 if SpO2 < 80%).

### 4.2 Implementation Protocol

The methodological framework for ORC testing in critically ill patients (including FiO2 titration logic and ORC calculation rules) aligns with the non-critical protocol. Its primary objective is to construct a dynamic, full-curve ORC profile across the entire disease course of critically ill patients, providing high-precision, continuous, and comprehensive quantitative data for studying infection-associated respiratory dysfunction. Due to the integration of high-risk procedures such as mechanical ventilation support and multi-organ monitoring in critical care settings, implementation must commence only after validation of the non-critical protocol. Respiratory support parameters, monitoring criteria, and safety protocols shall be strictly defined in an independent operations manual. Below is the complete implementation protocol for non-critical/early-stage testing.

#### 4.2.1 Subject Preparation

##### Inclusion Criteria

Adults (18–75 years) diagnosed with non-critical/early-stage respiratory infections (e.g., community-acquired pneumonia, early viral pneumonia).

Hemodynamic stability: systolic blood pressure ≥90 mmHg, heart rate ≤100 bpm; no acute respiratory distress (respiratory rate ≤24 bpm) or hypoxemia (resting SpO2 ≥94%).

##### Exclusion Criteria

Severe cardiopulmonary comorbidities, chronic respiratory failure, impaired consciousness, or inability to comply with testing.

##### Pre-Test Preparation

Positioning: Seated rest for ≥10 minutes; no physical activity, eating, or drinking to minimize respiratory variability.

Standardized Instructions: “Maintain sitting and natural breathing patterns; avoid deliberate deep breaths or breath-holding; minimize fluctuations in respiratory rate and tidal volume throughout testing.”

Environmental Controls: Room temperature 22–25°C, humidity 40–60%, no strong light or noise interference.

#### 4.2.2 Equipment and Monitoring Configuration

##### Core Devices

Altitude hypoxia generator, normobaric chamber, gas mixer, gas analyzer, respiratory flowmeter, Medical-grade pulse oximeter, bedside blood gas analyzer (for PaO2), non-invasive ventilator with respiratory mechanics monitoring.

##### Monitoring Parameters

Real-time: SpO2 (target ≥90%), tidal volume (VT), respiratory rate (RR), minute ventilation (VE).

Ancillary: ECG (exclude arrhythmias), non-invasive blood pressure (ensure hemodynamic stability).

#### 4.2.3 Standardized Ventilation Control

##### Baseline Calibration

FiO2 = 21%: Record VT, VE, and RR over 5 minutes (average values used as baseline). When the tester first tested, the system generates real-time respiratory rate (RR) baseline-synchronized auditory cues based on continuous monitoring of the subject’s breathing pattern. This audio signal will be looped in a standardized protocol during subsequent testing phases, utilizing auditory biofeedback mechanisms to guide the subject in maintaining RR baseline stability, thereby ensuring respiratory rate fluctuations remain within ±2 bpm.

Alarm Thresholds: VT fluctuation ≤±15%, VE ≤±20%, RR ≤±2 bpm.

##### Abnormal Protocol

If one of VT, VE, or RR exceeds the threshold for more than one minute: pause testing, troubleshoot (e.g., poor compliance), recalibrate, and re-establish stable baseline (VT fluctuation ≤±15%, VE ≤±20%, RR ≤±2 bpm for 2 minutes).

#### 4.2.4 FiO2 Titration Protocol

Initial Phase (Baseline):

FiO2 = 21% for 5 minutes; confirm SpO2 ≥94% and stable VT, VE, and RR.

##### Reducing Oxygen Concentration Phase

Stepwise FiO2 reduction: 1% decrement per minute (21% → 20% → 19%…) until SpO2

≤90%.

##### Termination Criteria

SpO2 <85%.

One of VT, VE, or RR exceeds the threshold for more than one minute. FiO2 <15%.

Emergency Response: Immediately restore FiO2 to 21% until SpO2 ≥94%.

#### 4.2.5 Data Acquisition and Calculation

##### Key Parameters

FiO2-MIN: Minimum FiO2 maintaining SpO2 ≥90% (precision: 0.1%).

ORC Calculation: ORC = 0.21–FiO2-MIN (e.g., FiO2-MIN = 0.15 → ORC = 0.06). Stability Annotation: Document VT, VE, and RR fluctuation ranges (e.g., “ORC = 0.06, VT ±4%”).

##### Dataset Construction

###### ➀ Acute Infection Phase

Testing Frequency: Conduct daily measurements within a fixed time window (e.g., 9:00–11:00) to minimize circadian rhythm interference, with continuous monitoring for ≥3 days. Adjust the duration based on individual recovery trajectories until Oxygen Reserve Capacity (ORC) stabilizes at baseline levels.

###### ➁ Chronic Phase (Analogous to Long COVID)

Testing Frequency: Perform standardized weekly or extended-interval monitoring (e.g., biweekly) until ORC returns to baseline.

Storage Format: Structured electronic spreadsheets (timestamp, FiO2-MIN, ORC, VT, RR, VE, SpO2 trends).

#### 4.2.6 ORC-β/γ Kinetic Modeling

##### ➀ORC Curve

Obtain the ORC values of the patient throughout the entire course of the disease through continuous monitoring, and fit the time ORC curve.

Figure 1 is the line chart of ORC simulation data of mild and severe respiratory tract infection, and Figure 2 is the line chart of ORC simulation data of non functional patients (finally recovered) of long-COVID syndrome, where time=0 represents the infection day.

**figure 1.**
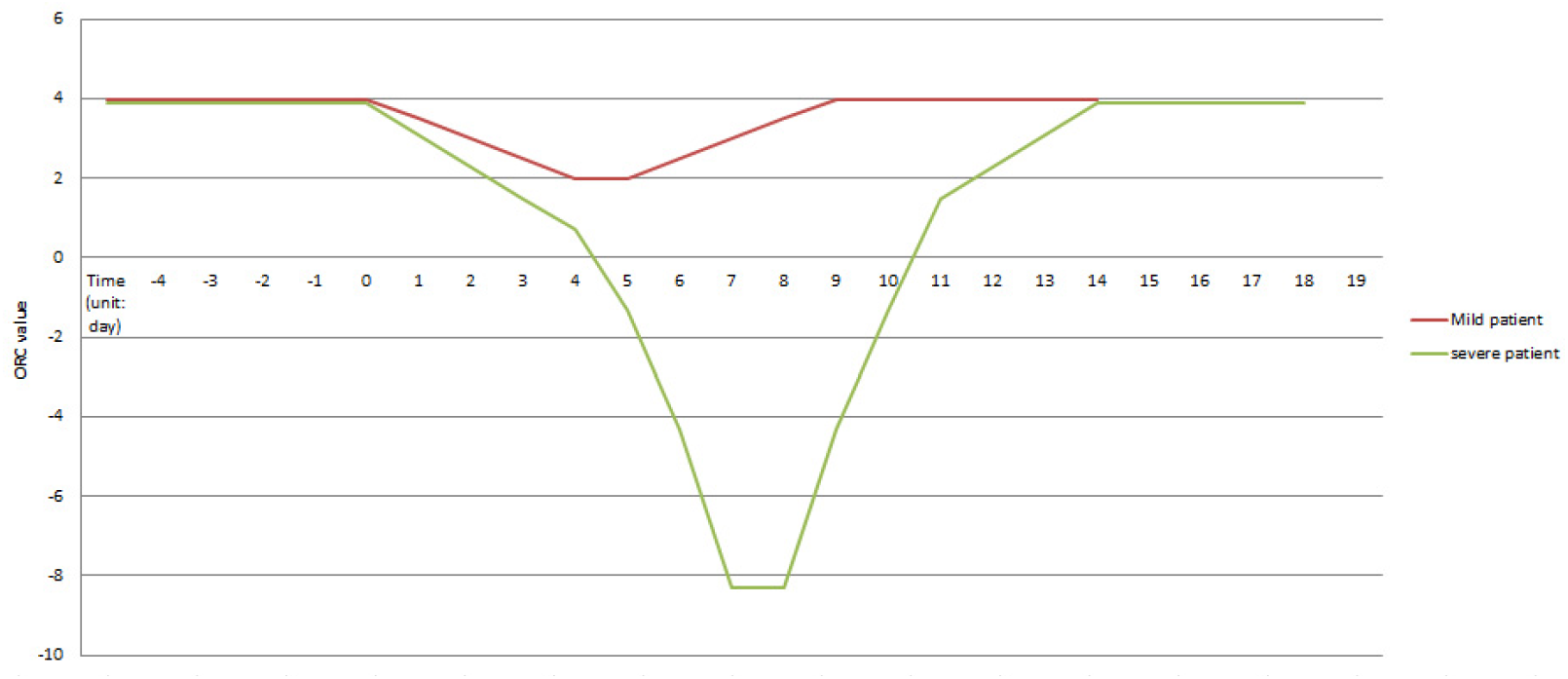
ORC simulation data of mild and severe respiratory tract infections.

**figure 2.**
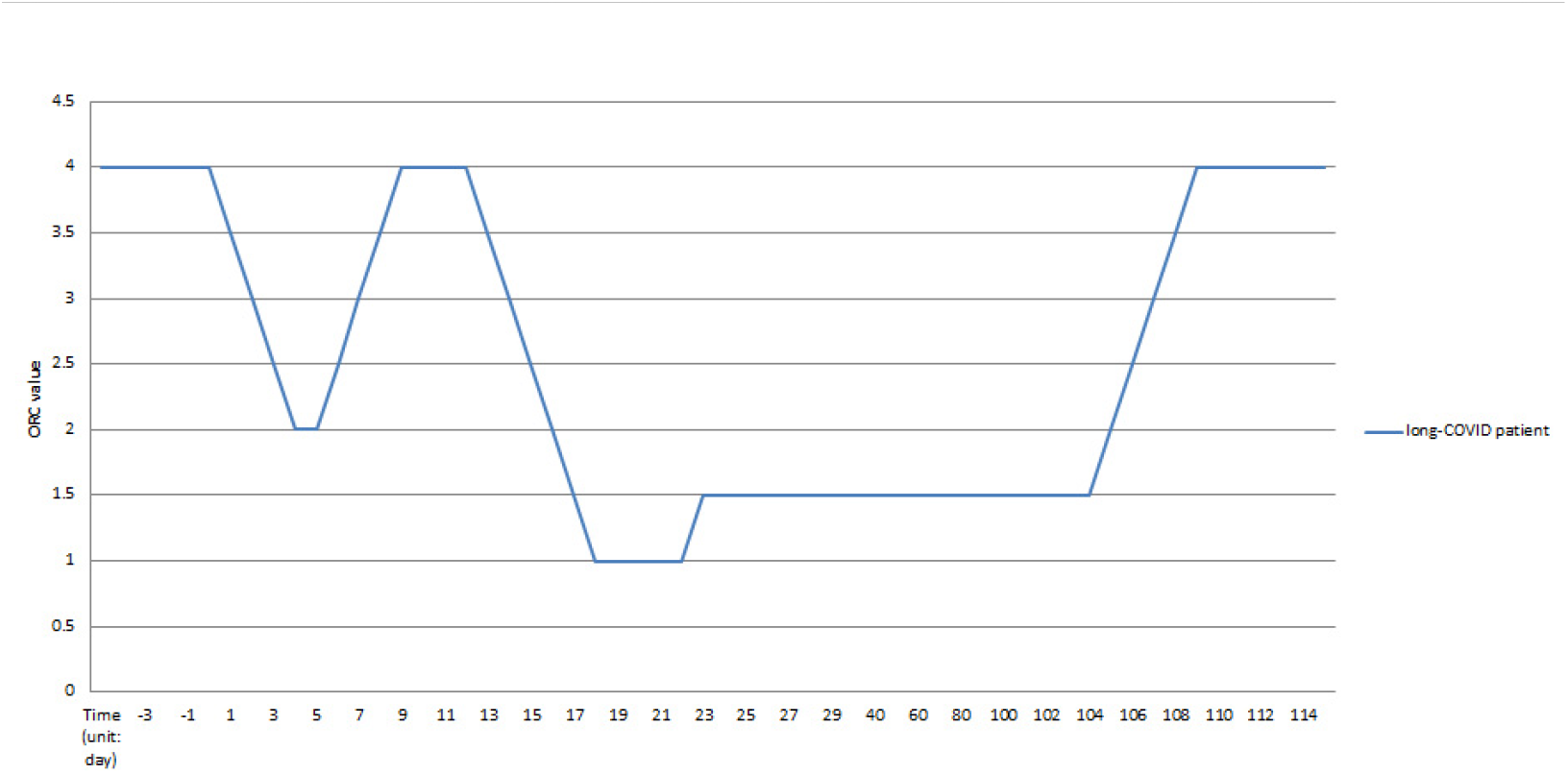
ORC simulation data chart of long-COVID.

➁β Parameter (Decay Rate of Compensation Efficiency):

Definition: β = ΔORC/Δt(unit: day−1), quantifying the decay rate of compensatory

efficiency over a 24-hour interval..

➂γ Parameter (Kinetics of Compensation Inflection):

Definition: γ=Δβ/Δt(unit: day−2), quantifying the turning point of respiratory compensation efficiency.

#### 4.2.7 Quality Control and Standardization

Operator Training:

Unified manual: device calibration (pre-use daily), standardized patient instructions, error code classification (e.g., E01: device failure; E02: non-compliance).

Data Validation:

Repeatability: Triplicate same-day measurements(excluding acute deterioration phases) (FiO2-MIN coefficient of variation, CV <5%).

Multi-Center Consistency: Regular cross-calibration of FiO2 sensors.

#### 4.2.8 Safety and Ethics

Emergency Protocols:

Immediate access to rescue equipment (endotracheal intubation kit, emergency medications).

Ethical Compliance:

IRB approval and informed consent (explicitly state risks and right to withdraw).

## 5. Discussion

### 5.1 Potential Clinical Applications

#### 5.1.1 Early Warning of ARDS Transition in Acute Infection

The ORC-β/γ kinetic model identifies “functional compensation blind spots” missed by traditional metrics (e.g., PaO2/FiO2 ratio). For instance, in early viral pneumonia, stable PaO2/FiO2 may mask significant ORC decline. This model could stratify ARDS transition risk, guiding timely interventions like prone positioning.

#### 5.1.2 Quantifying Long COVID Pulmonary Dysfunction

A subset of long COVID patients exhibit post-exertional oxygenation impairment (e.g., reduced VO2peak). The dynamic ORC test, based on a stepwise FiO2 titration protocol, quantifies pulmonary oxygen compensation thresholds (ORC = 0.21 − FiO2-MIN) under resting conditions through standardized ventilatory control and incremental FiO2 reduction. This approach circumvents limitations of cardiopulmonary exercise testing (CPET), such as exercise dependency, participant compliance variability, and equipment constraints. Accurate quantification and long-term dynamic tracking of non-invasive abnormal pulmonary function (ORC) can be achieved through Orc curve.

#### 5.1.3 Multidimensional Evaluation of Respiratory Infection Intervention Efficacy

Traditional efficacy assessment primarily relies on symptom scoring or imaging improvements. However, the ORC test provides complementary multidimensional assessments, such as the following:

Anti-Inflammatory Treatment Response

The ORC recovery rate (γ), compared to baseline values, quantifies the repair efficiency of the alveolar-capillary membrane during anti-inflammatory therapy.

#### 5.1.4 Unified Quantification of Prognostic Factors

The ORC test systematically analyzes multidimensional risk factors in respiratory infections—such as age, underlying comorbidities (e.g., diabetic microangiopathy), and environmental exposures (e.g., PM2.5)—and their distinct mechanistic impacts on ORC.

### 5.2 Limitations of ORC Testing

#### 5.2.1 Gaps in Practical Validation of Theoretical Models

The stepwise FiO2 titration protocol proposed in this study remains a theoretical framework and testing protocol design, lacking validation through large-scale clinical practice. Key unresolved issues requiring further exploration via multicenter clinical trials include:

➀Pathophysiological Relevance of ORC

Multimodal functional assessments (e.g., CT pulmonary perfusion imaging) must validate whether ORC decay truly reflects diffusion impairment or microcirculatory

decompensation.

➁Safety Threshold Controversies

The SpO2

#### 5.2.2 Challenges in Standardized Ventilation for Spontaneously Breathing Patients

➀Failure of Voice-Prompt-Controlled Respiratory Rate (RR) Standardization:

Standardized RR targets (e.g., 12-18 breaths/min) may lose efficacy due to fluctuations in patient consciousness or fatigue, leading to tachypnea (e.g., RR >25 breaths/min) or hypoventilation (e.g., VT

➁Real-Time Feedback Technology Limitations:

Requirements for standardized inspiratory/expiratory effort (via flow monitoring) may introduce errors from poor patient compliance (e.g., inconsistent tidal volume delivery) or device insensitivity (e.g., flow signal delays >200 ms).

To address the aforementioned challenges, it is recommended to prioritize the inclusion of subjects with high compliance into the research cohort, systematically implement standardized training protocols for tidal volume (VT) and respiratory rate (RR), and rigorously validate the validity and reliability of OCR testing under strictly controlled clinical operating procedures.

#### 5.2.3 Challenges in Baseline Data Integrity

The current ORC dynamic curve-based study design requires pre-infection individualized baseline values as reference standards to precisely quantify the magnitude of pulmonary oxygen reserve decay and recovery trajectories during infection. However, in clinical practice, patients often present post-acute infection without prior baseline data. To address this limitation, future strategies should focus on:

➀Promoting Non-Invasive Dynamic Monitoring: Popularize regular Orc testing on a large scale, build a longitudinal Orc database, and realize the continuous tracking of individualized baseline trends.

➁Prospective Data Collection: Implementing multicenter prospective cohort studies in high-risk populations (e.g., pre-infection baseline screening) to establish standardized, individualized ORC baseline datasets.

➂AI-Driven Baseline Prediction Models: Developing models using demographic data (age, BMI) and accessible physiological parameters, with cross-validation and external cohort testing to enhance generalizability.

#### 5.2.4 Technical Implementation Complexity

The ORC test requires high-precision hypoxic generators and respiratory mechanics monitoring equipment, posing cost and operational barriers for primary care facilities. For non-severe/early-stage patients, the following simplified strategies may be utilized to assess pulmonary oxygen reserve capacity:

➀Pulmonary oxygen reserve capacity may be assessed by adjusting nitrogen inhalation flow rates to modulate inspired oxygen concentration (FiO2), thereby simulating hypoxic conditions while using nitrogen concentration gradients to quantify changes in pulmonary oxygen reserves. The maximum nitrogen flow rate that maintains SpO2 ≥90% serves as a critical indicator to quantify pulmonary oxygen reserve capacity.

➁Controlled intermittent breath-holding protocols should be implemented to induce progressive hypoxic stimulation, with concurrent monitoring of physiological tolerance thresholds.

### 5.3 Future Research Directions

➀Multicenter Validation and Standardization

Cross-regional trials (incorporating diverse altitudes/climates) are needed to validate ORC universality and establish unified protocols (e.g., FiO2 decrement rates, SpO2 safety thresholds).

➁Integration with Multi-Omics Profiling

Combining proteomic (e.g., autoantibody-targeted panels) and ORC dynamic data could unravel molecular networks driving oxygen reserve decay, advancing precision medicine. 6.Conclusion

The stepwise FiO2 titration protocol, by dynamically profiling pulmonary oxygen reserve capacity (ORC), overcomes functional limitations of traditional tools, offering an innovative framework for studying infection-related respiratory dysfunction. However, clinical translation requires addressing baseline data gaps, technical standardization, and mechanistic validation. Future efforts must prioritize multicenter trials, device simplification, and molecular correlation studies to bridge the gap between theoretical models and real-world practice.

## 7. PS

The authors acknowledge that the interdisciplinary nature of this study may have led to oversimplifications in certain technical discussions. While rigorous peer validation was applied to core methodologies (e.g., ORC protocol design), the depth of domain-specific expertise (particularly in exercise-induced metabolic compensation mechanisms) could be further strengthened through collaborative research with sports physiologists in future work.

## Data Availability

All data produced in the present work are contained in the manuscript

